# The STRONG Study: A Multi-Tiered, Multimodal Investigation of Resilience and Recovery Following Prolonged Collective Adversity

**DOI:** 10.64898/2026.08.17.26360579

**Authors:** Dalia Moallem, Rinatia Maaravi-Hesseg, Daniel Panitz, Robert H. Pietrzak, Ziv Ben-Zion

## Abstract

Stress-related disorders are among the most common and burdensome mental health conditions worldwide, yet the mechanisms that allow most trauma-exposed individuals to maintain or regain mental health remain poorly understood. Decades of research have focused on identifying risk factors for psychopathology rather than the active processes that promote resilience and recovery. Here, we present the study protocol for **Stress and Trauma Resilience: Opportunities for National Growth (STRONG)** — a multi-tiered, multi-domain, multi-level investigation of resilience conducted in Israel in the aftermath of the October 7, 2023 attack and the prolonged national adversity that followed. STRONG uses a nested design that integrates nationally representative longitudinal data with in-depth neurobehavioral assessment. STRONG-1 is a longitudinal, population-based study of approximately 4,600 Israeli adults assessed across five waves over three years, characterizing individual, social, and societal predictors of resilience trajectories. STRONG-2 is a controlled laboratory study of highly resilient and highly vulnerable individuals selected from STRONG-1, assessing behavioral and physiological mechanisms alongside cognitive tests and ecological momentary assessment. STRONG-3 examines a subset of these individuals in the MRI scanner, capturing structural and functional neural markers with synchronized physiological and eye-tracking data. Advanced computational approaches will integrate data across tiers, levels, and domains into predictive models of resilience. STRONG will establish Israel’s first nationally representative dataset on stress resilience and provide a rare opportunity to study human adaptation at scale and in a real-world context. These findings will inform early detection strategies and the development of empirically grounded, modifiable targets for intervention.

**Preprint status:** This is a living protocol. STRONG-1 (the population-based tier) has completed its baseline (T1) assessment and is described here at the level of what was administered. STRONG-2 and STRONG-3 (the laboratory-based and neuroimaging tiers), and the selection procedure that links them to STRONG-1, are still being finalized and are described here at the level of design and planned measures; specifics may change in subsequent versions and in the peer-reviewed version of record. Preregistered hypotheses, analysis plans, materials, and code are — or will be — available on the Open Science Framework (OSF): https://osf.io/r56pb

## 1. Background and Rationale

On October 7, 2023, Israel experienced the deadliest attack on Jewish people since the Holocaust. What followed was not a single, bounded event but prolonged national adversity (war, forced displacement, repeated alarms, and pervasive uncertainty) that continues to shape daily life. Under this shared burden, individuals and communities have diverged sharply. Some rebuild through social connection, adaptive coping, and renewed purpose; others remain in cycles of chronic anxiety, posttraumatic stress, and disrupted functioning. What distinguishes those who maintain or regain well-being from those who remain vulnerable? This question is the central motivation for the present work.

### 1.1 The paradox of trauma research

Trauma exposure is a prerequisite for posttraumatic stress disorder (PTSD)^1^, yet most trauma-exposed individuals do not develop chronic psychopathology. Large-scale longitudinal studies consistently show that the majority either maintain stable mental health or recover relatively quickly^2,3^. Historically, however, trauma research has concentrated on risk factors for psychopathology rather than the natural mechanisms that promote resilience and recovery in most people. This emphasis is understandable, but it is also limiting. As large-scale stressors — pandemics, economic crises, wars, and disasters — become more frequent, there is a pressing need to shift from a pathology-centered account to one that treats resilience as an active, adaptive, and dynamic process^4^.

### 1.2 Conceptualizing resilience as a dynamic, multi-domain, multi-level process

Borrowed from material science, “resilience” originally described the capacity to “bend and not break.” Early work framed it as a stable personality trait, but this view failed to identify a single robust predictor of mental health across trauma-exposed populations^4,5^. Contemporary frameworks instead define resilience as a dynamic process of adaptation shaped by multiple fluctuating and interacting factors^6,7^. A growing consensus operationalizes resilience as a positive mental health outcome, achieved either by maintaining functioning, or rapid return to baseline, despite significant adversity^8^. We adopt “resilience” to encompass both definitions, and use “recovery” when referring specifically to those who experience initial distress but later regain well-being.

Resilience is not just an outcome but a process that unfolds over time. Longitudinal studies using latent growth mixture modeling (LGMM)^9^ have repeatedly revealed four common post-trauma trajectories^2^: resilience (stable low distress), recovery (initial distress followed by return to low symptoms), chronic distress (persistent high symptoms), and delayed onset (symptoms that emerge over time). These patterns have been replicated across disaster survivors, military personnel, and trauma-exposed civilians. In our own prior work following 100 high-risk survivors diagnosed with PTSD one month after trauma, 71% naturally recovered by 14 months despite their initial vulnerability^10^. Recovery, in other words, is common even among highly symptomatic individuals — and it is the mechanisms underlying these divergent trajectories that STRONG is designed to uncover.

Resilience emerges from processes operating across distinct domains and contextual levels. At the individual level, STRONG focuses on five core domains: psychological, cognitive, behavioral, physiological, and neural. Each domain is independently linked to adaptive outcomes^11^. These domains have too often been studied in isolation, obscuring their combined and interacting influence. They are also embedded within a broader social level (e.g., perceived support, family cohesion) and societal level (e.g., perceived safety, institutional trust, access to natural spaces), which are rarely examined alongside neurobehavioral factors^12^. A recent systematic review identified four converging gaps: a limited range of studied resilience factors, insufficient attention to social and societal variables, a lack of cross-level modeling, and inadequate methods for capturing temporal dynamics^12^. STRONG is built to address these gaps directly.

### 1.3 Key gaps STRONG is designed to fill

Most resilience research has examined isolated domains with limited integration. Studies typically focus either on large-scale epidemiological cohorts or on controlled laboratory paradigms, limiting ecological validity or mechanistic insight, respectively. Cross-sectional and short-term designs dominate, failing to capture resilience as a dynamic process. Social and societal factors remain underexplored and are seldom modeled alongside individual ones. Trauma research has largely focused on single-event exposures, despite the reality that most people face cumulative and ongoing stress. And neurobiological studies have disproportionately targeted vulnerability rather than the mechanisms of resilience and recovery. STRONG confronts these limitations through a tiered, nested design that bridges broad population-based insight with in-depth neurobehavioral assessment, clarifying not only who is resilient but how and why resilience emerges.

### 1.4 The present protocol

Here we present the protocol for STRONG, a program combining psychology, neuroscience, and public health within a unified multi-domain, multi-level framework. STRONG will be the first nationally representative resilience study in Israel and, to our knowledge, the first to integrate population-based trajectory modeling with laboratory-based behavioral, physiological, and neural assessment following a real-world collective trauma. We describe the completed baseline of the population tier (STRONG-1) in detail, and the planned laboratory (STRONG-2) and neuroimaging (STRONG-3) tiers at the design and candidate-measure level.

## 2. Aims and Objectives

STRONG follows a three-tiered, nested design of progressively increasing depth and precision (Figure 1). STRONG-1 identifies individual (e.g., psychological, physical health and lifestyle), social, and societal predictors in a nationally representative cohort (Figure 2); STRONG-2 assesses behavioral, cognitive, and physiological mechanisms in a laboratory setting; and STRONG-3 identifies structural and functional brain correlates. The program is organized around four aims. Specific confirmatory, directional hypotheses and analysis plans for individual studies will be specified in separate OSF preregistrations.

**Figure 1.**
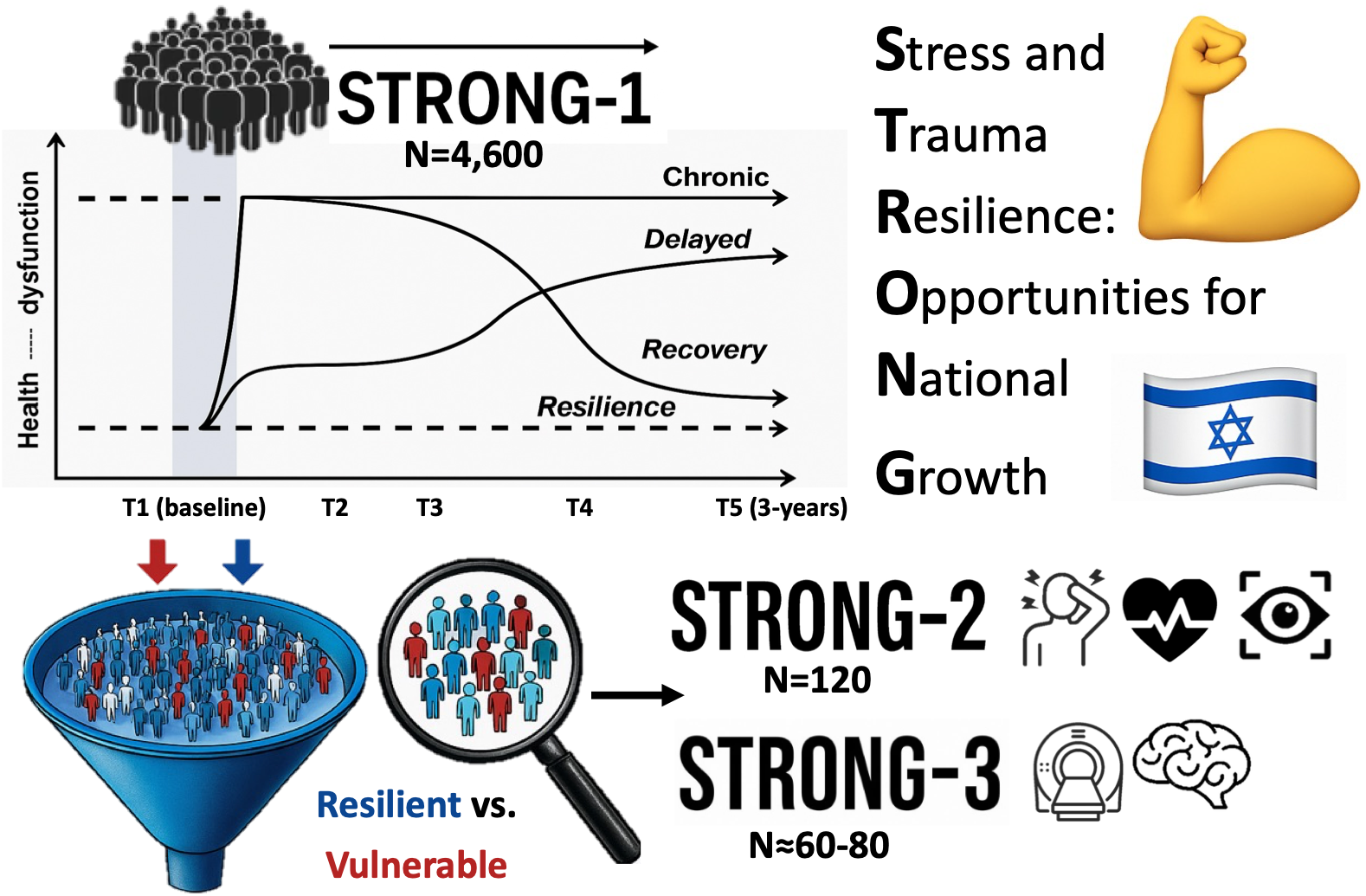
The STRONG Study Overview. A visual summary of STRONG’s three-tiered, nested design, illustrating the progression from population-based trajectory modeling (STRONG-1) to mechanistic behavioral (STRONG-2), and neuroimaging (STRONG-3) assessments.

**Figure 2.**
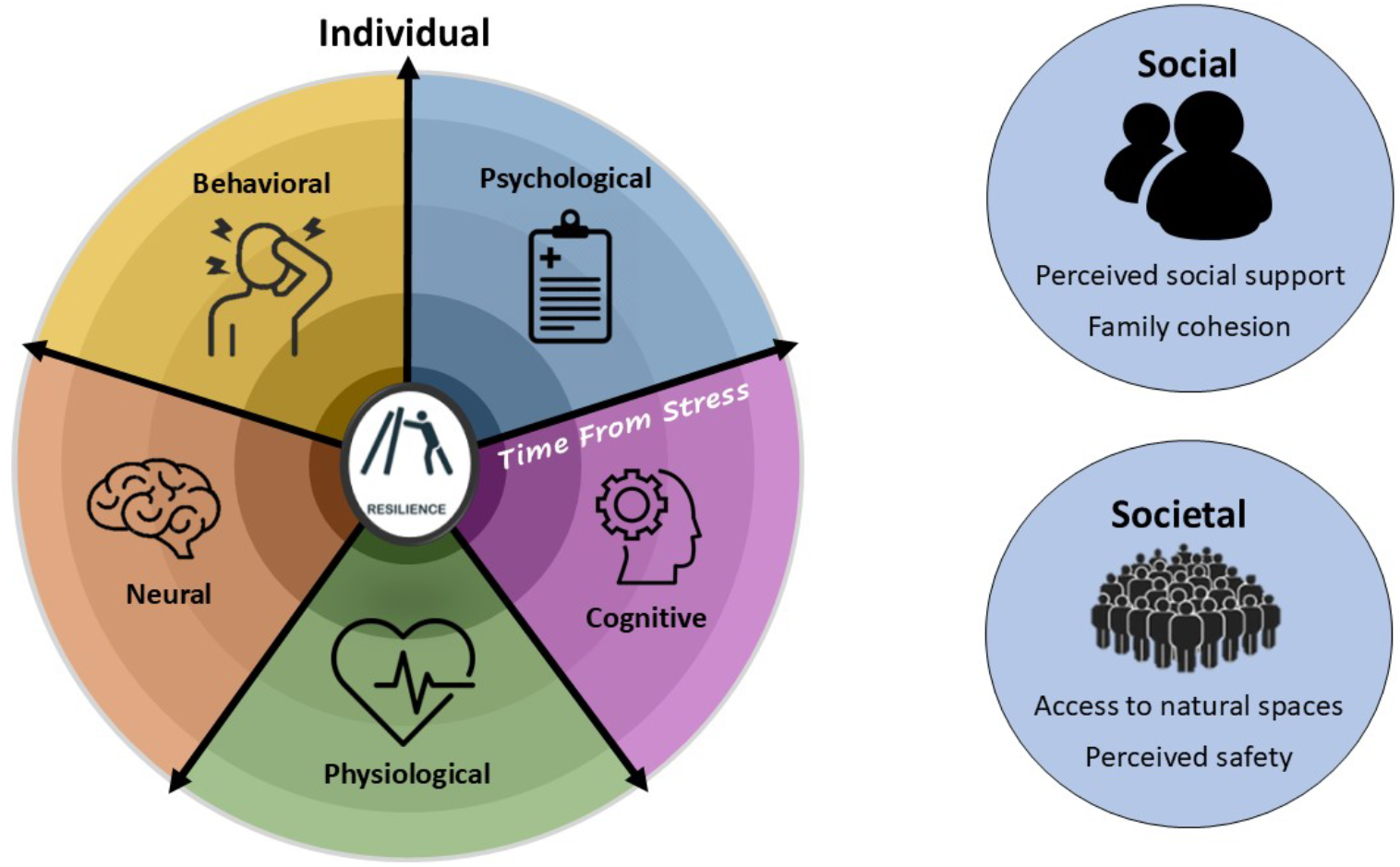
The STRONG Study Multi-Domain, Multi-Level Framework. Stress resilience emerges from the dynamic interaction of processes across five individual-level domains—psychological, cognitive, physiological, behavioral and neural—and their broader social and societal contexts.

### Aim 1 — Identify individual, social, and societal correlates of resilience

Using STRONG-1 baseline data, we will: (1.a) estimate the prevalence of resilience in the Israeli population following prolonged national adversity; (1.b) identify individual-level demographic, physical health, lifestyle, and psychological correlates; (1.c) identify social and societal predictors; (1.d) examine cross-level interactions; and (1.e) compare findings to international resilience studies (e.g., the U.S. National Health and Resilience in Veterans Study, NHRVS^13^) to distinguish universal from context-specific factors. We will estimate the prevalence of high resilience despite ongoing adversity, and examine whether resilience is associated with positive appraisal style, grit, self-compassion, and emotion regulation (1.b); with higher perceived social support, institutional trust, and perceived safety (1.c); and whether social and societal resources buffer lower psychological capacities (1.d).

### Aim 2 — Identify behavioral, physiological, and neural markers of resilience

Using STRONG-2 and STRONG-3, we will contrast highly resilient and highly vulnerable individuals selected from STRONG-1 on: (2.a) cognitive, behavioral, and physiological responses during experimental tasks; (2.b) structural and functional brain measures; (2.c) real-world emotion regulation, affect, and coping via ecological momentary assessment (EMA); and (2.d) converging multi-domain mechanisms. We will examine whether resilient individuals will show more adaptive physiological responses to acute stress (e.g., higher baseline heart rate variability) and more adaptive behavioral patterns (e.g., greater reward sensitivity); whether neural correlates of resilience include larger hippocampal volume, enhanced prefrontal–amygdala connectivity, greater responsivity toward rewards versus punishments, and altered large-scale network connectivity; and whether adaptive markers co-occur across domains (e.g., a “resilient reward profile” spanning behavior, physiology, and brain).

### Aim 3 — Identify dynamic predictors and mechanisms of resilience trajectories

Using STRONG-1 longitudinal data (T1–T5), we will: (3.a) identify latent trajectories of mental health outcomes via LGMM; (3.b) assess how baseline levels and temporal changes in individual, social, and societal factors predict these trajectories; and (3.c) test whether neurobehavioral markers from Aim 2 predict trajectory membership. We will test whether the four common trajectories replicate (resilience, recovery, chronic distress, delayed onset); whether adaptive trajectories are predicted by higher baseline levels — and positive changes over time — in measured resilience-relevant constructs (e.g., positive appraisal, grit, self-compassion, emotion regulation), especially when supported by social and societal resources; and whether neurobehavioral markers differentiate resilience and recovery trajectories beyond self-report.

### Aim 4 — Develop a multi-domain, multi-level predictive model of resilience

Using advanced statistical and machine-learning approaches, we will integrate data across STRONG-1, -2, and -3 to (1) optimize prediction of resilience and recovery trajectories and (2) identify the most influential and modifiable predictors. These interpretable models will clarify how predictors interact across domains and levels, highlight compensatory pathways, and point toward mechanism-based intervention targets.

## 3. Methods

The STRONG project employs a three-tiered, nested design (Figure 1), progressing from broad population-based prediction (STRONG-1) to laboratory-based behavioral (STRONG-2) and neural (STRONG-3) assessment (Figure 3). This structure yields both breadth and mechanistic precision, and it allows the same well-characterized individuals to be followed across levels of analysis.

**Figure 3.**
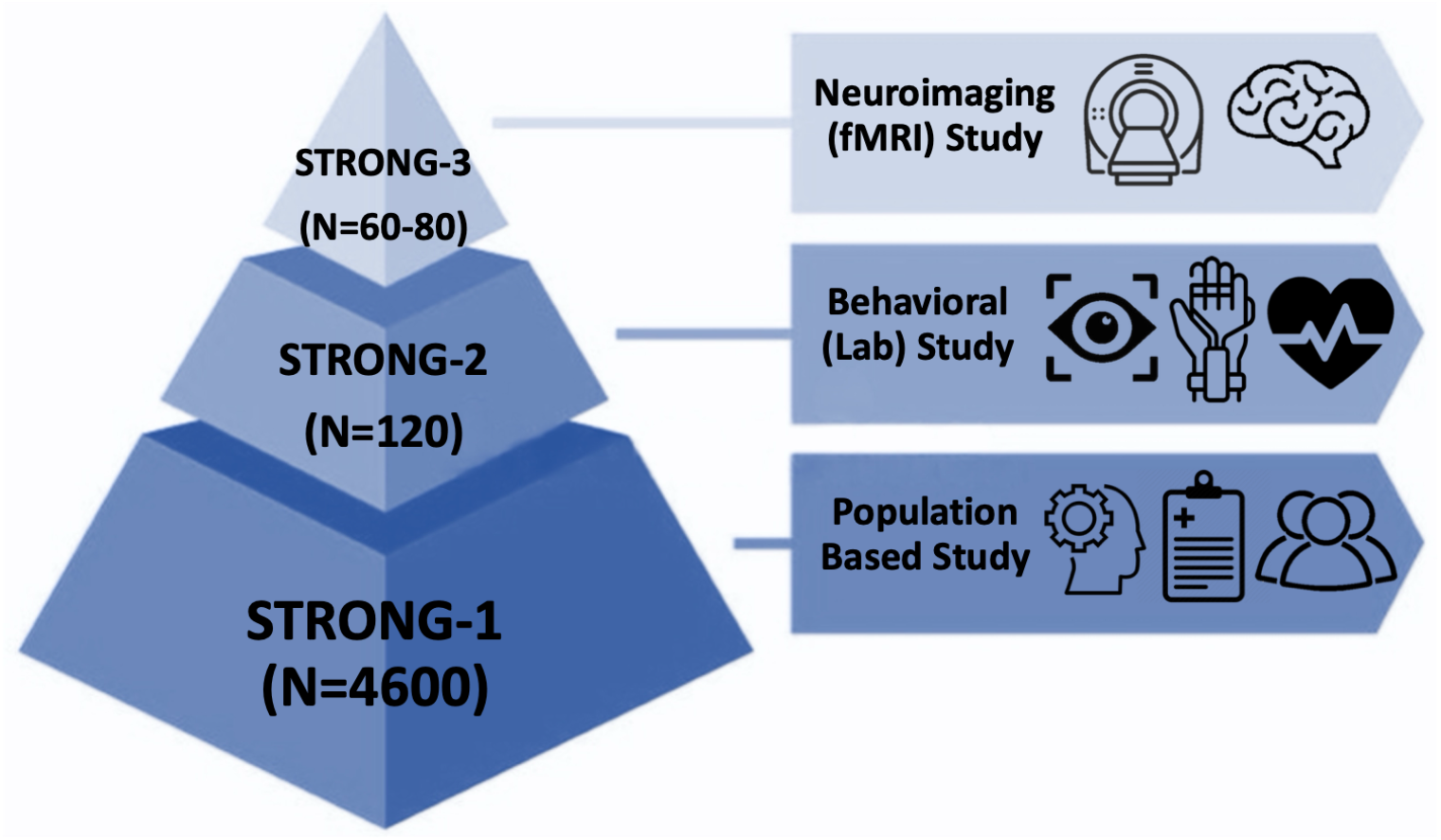
Tiered Structure of the STRONG Study. A three-tiered, nested design integrates population-based (STRONG-1), behavioral (STRONG-2), and neuroimaging (STRONG-3) assessments to investigate resilience across domains and levels.

### 3.1 STRONG-1: Population-Based Longitudinal Study

#### Status

STRONG-1 baseline (T1) data collection is complete. The description below reflects the assessment as administered; longitudinal waves (T2–T5) are planned.

#### Participants and procedure

STRONG-1 is a longitudinal, national study of resilience and vulnerability trajectories and their individual, social, and societal predictors. Self-reports from approximately 4,600 Israeli adults (aged 18+) are assessed at five time points over three years (T1–T5: baseline, 6 months, 1 year, 2 years, 3 years), capturing long-term mental health trajectories. Participants are recruited and monitored through Panel4All, an online Israeli panel company with over 100,000 Israelis in their database. Quota sampling and post-sampling weighting align the sample with Israeli census data across age, gender, ethnicity, religion, education level, and geographic location. Recruitment uses online methods, with accommodations for Hebrew, Arabic, and English speakers. Retention is supported through personalized follow-ups, financial incentives, and evidence-based engagement protocols. Based on prior longitudinal work of a similar nature using Panel4All^14^, we anticipate that at least 60% of participants will complete all waves, ensuring sufficient power (Section 4). Wave-by-wave data permit evaluation of attrition bias and trajectory modeling. At baseline, participants also indicated willingness to take part in follow-up studies (STRONG-2 and STRONG-3), supporting the nested design.

#### Measures

The STRONG-1 battery spans individual (demographic, psychological), social, and societal domains, along with stressors specific to the October 7, 2023 attack and subsequent war. Many of these self-report measures are validated and the rest were created and/or adapted by the study team to be appropriate for the current situation in Israel (see Supplementary Table S1 for the full list of constructs, measures, number of items, and citations).

- **Demographics**. Age, gender, religion, socioeconomic status (SES), education, marital/family status, income, and geographic location.
- **Psychological — traumatic exposure**. Lifetime trauma exposure was measured with the Life Events Checklist for DSM-5 (LEC-5^15^) and the Adverse Childhood Experiences scale (ACE^16^), and childhood unpredictability with the brief Questionnaire of Unpredictability in Childhood (QUIC-5^17^). Post–October 7 exposures (e.g., forced displacement, bereavement, reserve duty, moral injury, media exposure) were assessed with an investigator-developed October 7/Israel-specific questionnaire and, for those with relevant service history, an investigator-developed Army Service questionnaire — included given their salience to the present population and moment.
- **Psychological — symptoms**. PTSD symptoms (PTSD Checklist for DSM-5, PCL-5^18^), depression (Patient Health Questionnaire-9, PHQ-9^19^), anxiety (Generalized Anxiety Disorder-7, GAD-7^20^), sleep difficulties (brief Pittsburgh Sleep Quality Index, PSQI^21^), suicidality (Columbia-Suicide Severity Rating Scale, C-SSRS^22^), perceived stress (Perceived Stress Scale, PSS-10^23^), obsessive-compulsive symptoms (4-Item Obsessive-Compulsive Inventory, OCI-4^24^), and substance use (ASSIST-Lite^25^). Current diagnoses, treatment status, and use of AI for emotional support were assessed with investigator-developed items.
- **Psychological — resilience-relevant constructs**. Trait resilience was measured with the 10-item Connor-Davidson Resilience Scale (CD-RISC-10^26^, adapted from Connor & Davidson^27^) and the 24-item Mount Sinai Resilience Scale (MSRS^28^). Additional constructs included personality traits (Ten-Item Personality Inventory, TIPI^29^), emotion regulation (Emotion Regulation Questionnaire – Short Form, ERQ-S^30^), positive appraisal style (Positive Appraisal Style Scale – Content, PASS-Content^31^), grit (Short Grit Scale, Grit-S^32^), self-compassion (Sussex-Oxford Compassion for the Self Scale, SOCS-S^33^), optimism (Life Orientation Test, LOT^34^), religiosity (The Duke University Religion Index, DUREL^35^), posttraumatic growth (Posttraumatic Growth Inventory, PTGI^36^), and functional participation in routine activities (Oxford Participation and Activities Questionnaire – Routine Activities, OxPAQ-RA^37^).
- **Social level**. Social support (modified Medical Outcomes Study Social Support Survey, mMOS-SS^38^), loneliness (3-item UCLA Loneliness Scale^39^), and community resilience (Conjoint Community Resiliency Assessment Measure, CCRAM^40^).
- **Societal level**. National resilience, including items assessing institutional trust (National Resilience Scale^41^), and perceived sense of danger (Sense of Danger Scale; adapted from Kimhi & Eshel’s national resilience research^42^).
- **Physical health and lifestyle**. Preexisting conditions, height, weight, physical activity (International Physical Activity Questionnaire – Short Form, IPAQ-S^43^) and food-related intrusive thoughts (Food Noise Questionnaire, FNQ^44^).

#### Data analysis

Resilience is operationalized using a discrepancy-based psychiatric resilience (DBPR) approach^45^ — originally developed by Amstadter and colleagues^46^ and applied within the NHRVS^47^ — in which resilience is defined as better-than-expected mental health given an individual’s cumulative adversity. Specifically, DBPR will be estimated by regressing mental health outcomes using a composite distress score (PCL-5, PHQ-9 and GAD-7) on a cumulative adversity index; residuals reflect the gap between observed and model-predicted symptom levels, with better-than-predicted functioning (i.e., fewer symptoms than adversity exposure would predict) indicating resilience. The adversity index will combine LEC-5, ACE, October 7–specific exposure, and Army service exposures. This approach ensures that resilience reflects adaptive functioning rather than low exposure. We will estimate the reliability of DBPR scores (e.g., split-half/bootstrap) and verify their approximate independence from raw adversity exposure. In addition to DBPR, we will take into account function (utilizing OxPAQ-RA) and self reports of resilience (using CD-RISC-10 and MSRS) to categorize participants as resilient. LGMM will identify distinct symptom trajectories; multivariate regressions will relate these to baseline individual, social, and societal factors; and cross-level interactions will be examined via moderation and mediation. Missing-data handling will be specified per analysis, using approaches appropriate to each study. All procedures comply with applicable data-protection standards (Section 5).

### 3.2 STRONG-2: Laboratory-Based Behavioral and Physiological Study

#### Status

Planned. Participant selection and the final task battery are being finalized; the description below is at the design and candidate-measure level.

#### Participants and procedure

STRONG-2 is a controlled laboratory study examining psychological, cognitive, behavioral and physiological resilience markers. A subset of approximately 120 participants will be selected from STRONG-1 based on baseline DBPR scores and accompanying resilience measures: roughly 60 highly resilient and 60 highly vulnerable individuals. The exact decision rule (e.g., a composite score, required agreement across measures, or another approach) has not yet been finalized. Groups will be matched on key variables (e.g., age, gender, SES, lifetime trauma). Because these variables are retained for all STRONG-1 participants, selection bias can be evaluated directly.

#### Candidate measures

STRONG-2 participants are expected to repeat the STRONG-1 self-report, complete mobile cognitive batteries and ecological momentary assessments, undergo clinical interviews to assess resilience and coping strategies, and perform validated experimental tasks targeting core behavioral resilience mechanisms (candidate paradigms include acute stress induction, reward processing, positive appraisal, and cognitive flexibility). Physiological markers at rest and during tasks will include electrocardiography and impedance cardiography (MindWARE Technologies BioNex), eye-tracking indices (Tobii Pro Fusion 250 Hz) and facial recognition features (Noldus Information Technology FaceReader Advanced). Participants are also expected to complete an at-home cognitive functioning assessment and EMA protocol^48,49^ capturing real-time fluctuations in stress, affect, and coping and their association with cognitive functioning.

#### Planned analysis

Group differences in behavioral, physiological, and EMA-based markers will be analyzed using appropriate statistical approaches (e.g., mixed-effects models). Cardiovascular measures — including indices derived from electrocardiography and impedance cardiography — will be used to characterize autonomic and hemodynamic reactivity and recovery (e.g., HRV, change scores, recovery slopes). Eye-tracking data will be analyzed using established gaze-dynamics approaches. EMA data will be modeled using approaches suited to nested, within-person data (e.g., hierarchical/multilevel models). Additional analytic approaches (e.g., profile-based methods such as latent profile analysis) may be used to characterize distinct resilience profiles and their associations with other study variables.

### 3.3 STRONG-3: Neuroimaging Study

#### Status

Planned. Participant selection and the final task battery are being finalized; the description below is at the design and candidate-measure level.

#### Participants and procedure

STRONG-2 participants will be invited to complete an MRI session at the University of Haifa Neuroimaging Research Unit (UH-NRU), equipped with a 3T scanner. Accounting for expected non-consent and MRI contraindications from the 120 STRONG-2 participants, we anticipate a final sample of approximately 60–80 (50–67%) across both resilience groups.

#### Candidate measures

Neuroimaging is expected to include high-resolution structural MRI, resting-state fMRI, and task-based fMRI. Candidate tasks include a risky decision-making paradigm probing anticipated rewards and punishments^50^, and a short emotionally evocative video. Concurrent MRI-compatible physiological and eye-tracking signals (HRV, GSR, pupil size, blink rate, fixation) will be collected to index arousal, attention, and regulatory effort in the scanner.

#### Planned analysis

Structural data will be processed with *FreeSurfer*. Resting-state and task-based fMRI will be preprocessed using *fMRIprep*. Preprocessed data will be analyzed (with *SPM* or *FSL*) for group differences in activation and connectivity using whole-brain and region-of-interest approaches. Physiological signals and eye-tracking measures will be synchronized with task events and modeled alongside fMRI data.

### 3.4 Integrative Multi-Domain, Multi-Level Predictive Modeling (Aim 4)

To address Aim 4, data from all three tiers will be integrated into a unified predictive model of dynamic resilience and recovery. Using advanced statistical and computational approaches, we will identify key predictors and mechanisms across domains and levels. Analyses will compare predictive accuracy across models, isolate the most influential and modifiable predictors, and characterize how these evolve over time to shape resilience and recovery trajectories.

## 4. Sample Size and Power

STRONG-1’s baseline sample (approximately 4,600 respondents who completed the T1 assessment) supports precise selection of well-characterized resilient and vulnerable subgroups for STRONG-2 (N ≈ 120) and STRONG-3 (N ≈ 60–80). Based on Panel4All’s longitudinal retention in a comparable study^14^ (≥ 60% over 2.5 years), STRONG-1 is expected to retain enough participants across all waves to provide ≥ 80% power to detect small effects (Cohen’s d ≈ 0.2; OR ≈ 1.5) in the population data. STRONG-2 and STRONG-3 are powered (≥ 80%) to detect small-to-medium effects (d ≈ 0.3–0.5; OR ≈ 1.5–2.5) in group comparisons. These estimates are grounded in comparable studies. Final sample sizes for STRONG-2 and STRONG-3 will be determined by funding and recruitment and reported in subsequent versions.

## 5. Ethics and Open Science

STRONG-1 was approved by the Ethics Committee, Faculty of Social Welfare and Health Sciences, University of Haifa (approval no. 045/26). Ethics approvals for STRONG-2 and STRONG-3 are in process. All STRONG-1 participants provided informed consent; participants in STRONG-2 and STRONG-3 will provide informed consent. Data handling will comply with applicable data security and privacy regulations, including secure storage, access controls, and de-identification.

Consistent with our commitment to transparent and reproducible science, where applicable, study hypotheses and analysis plans will be preregistered on the Open Science Framework (OSF). Given the program’s staged nature, any preregistration will proceed in tiers, with STRONG-2 and STRONG-3 registered as their designs are finalized. De-identified data and analysis code will be shared via public repositories to support transparency, replicability, and reuse.

## 6. Status and Timeline

STRONG-1 baseline (T1) data collection is complete (9 June – 9 July, 2026), and longitudinal follow-ups are scheduled across three years. Selection of STRONG-2 and 3 participants, the STRONG-2 laboratory battery, and the STRONG-3 neuroimaging protocol are being finalized. Subsequent versions of this protocol will report these details as they are locked, and the peer-reviewed version of record will be submitted once the full design is fixed.

## 7. Discussion

Stress and Trauma Resilience: Opportunities for National Growth (STRONG) addresses a pressing scientific and public health priority: understanding the mechanisms that promote resilience under prolonged stress. This protocol describes a program that complements a pathology-focused account with an adaptation-focused one and integrates psychology, neuroscience, and public health within a unified, multi-domain, multi-level framework. To our knowledge, STRONG is the first resilience study to combine nationally representative longitudinal data with targeted neurobehavioral assessment, linking population-based trajectories (STRONG-1) with behavioral (STRONG-2) and neural (STRONG-3) mechanisms, following an ongoing collective trauma.

Several design features distinguish STRONG. It treats resilience as a dynamic process, tracking mental health trajectories over time rather than at a single point. It jointly assesses five individual-level domains — psychological, cognitive, behavioral, physiological, and neural — many of which have rarely been studied together and never at a national scale, and it situates them within their social and societal contexts. At-home, mobile-based cognitive tasks complement in-lab assessments with dynamic measures of modifiable markers such as cognitive flexibility, further enhanced by real-time EMA^48^. The program also integrates leading resilience frameworks, namely regulatory flexibility and positive appraisal style, within a large, diverse population exposed to genuine adversity, using advanced analytics to integrate data across tiers, levels, and domains.

The program is ambitious, and its staged design carries real challenges. Integrating complex, multi-level, multi-domain data is nontrivial, and STRONG addresses this through a nested design and a clearly specified analytic pipeline using validated tools tailored to each data type. Retention across five waves is a well-known threat to longitudinal cohorts; it is mitigated by evidence-based engagement protocols and evaluated directly through wave-by-wave attrition modeling. Selecting laboratory subgroups from a population cohort risks selection bias; because key variables are retained for all STRONG-1 participants, this bias can be estimated rather than assumed away. Applying DBPR to ongoing, collective adversity rather than the discrete trauma histories on which it was originally developed is a further limitation, as the current adversity index reflects exposure to distinct event types rather than the frequency of repeated exposures (e.g., number of alarms or displacement episodes), which may understate cumulative burden for participants facing ongoing threat. Future waves or index refinements could incorporate frequency-weighted exposure to address this gap. Finally, the neurobehavioral tiers are not yet fully specified, which is precisely why this version of the protocol describes them at the level of design, to be refined in subsequent versions without overcommitting to details that may change.

Beyond its scientific contributions, STRONG will establish Israel’s first nationally representative dataset on stress resilience, a scalable platform for long-term mental health monitoring during and after large-scale adversity. By bridging psychology, neuroscience, and public health, and by identifying modifiable, mechanism-based targets across domains and levels, STRONG is designed not only to characterize who is resilient but to clarify how and why resilience emerges. This will hopefully open new avenues for early detection and for the development of scalable, personalized approaches to promoting resilience and recovery following traumatic stress.

## Data Availability

De-identified data and analysis code will be shared via public repositories to support transparency, replicability, and reuse.

