## Supplemental Table S1 for "The STRONG Study: A Multi-Tiered, Multimodal Investigation of Resilience and Recovery Following Prolonged Collective Adversity"

### Supplementary Table S1. Measures Included in STRONG-1

*Note. "Modified?" indicates whether the version used deviates from the original published measure (e.g., item selection, response scale, wording, or brevity). Items marked "N/A – custom" or "N/A – standard demographic items" are investigator-developed and have no external citation. Two rows are flagged (*) where the source data contained internal inconsistencies (item counts) that should be confirmed against the final survey before publication.*

| Construct | Measure Name | # Items | Citation | Modified? |
|---|---|---|---|---|
| **Traumatic Exposures** | | | | |
| Traumatic Events – Adulthood | Life Events Checklist for DSM-5 (LEC-5) | 17 | Weathers et al. (2013) | No |
| Traumatic Events – Childhood | Adverse Childhood Experiences (ACE) | 10 | Felitti et al. (1998) | No |
| Traumatic Events – Since Oct. 7, 2023 | October 7/Israel-Specific Questionnaire | 11 | Investigator-developed | N/A – custom |
| Traumatic Events – Since Oct. 7, 2023 (Army-specific) | Army Service Questionnaire | 8 | Investigator-developed | N/A – custom |
| Trauma/Stress – Childhood | Questionnaire of Unpredictability in Childhood – Brief (QUIC-5) | 5 | Lindert et al. (2022) | No |
| **Psychopathology** | | | | |
| PTSD | PTSD Checklist for DSM-5 (PCL-5) | 20 | Weathers et al. (2013) | No |
| Depression | Patient Health Questionnaire-9 (PHQ-9) | 9 | Kroenke, Spitzer, & Williams (2001) | No |
| Anxiety | Generalized Anxiety Disorder-7 (GAD-7) | 7 | Spitzer, Kroenke, Williams, & Löwe (2006) | No |
| Sleep | Brief Pittsburgh Sleep Quality Index (PSQI) | 6 | Buysse et al. (1989) | Yes – brief version (items 1–4, 5.2, 9 of full PSQI) |
| Suicidality | Columbia-Suicide Severity Rating Scale (C-SSRS) | 5 | Posner et al. (2011) | Yes- omitted items 3-6 |
| Perceived Stress | Perceived Stress Scale (PSS-10) | 10 | Cohen, Kamarck, & Mermelstein (1983) | No |
| OCD | 4-Item Obsessive-Compulsive Inventory (OCI-4) | 4 | Abramovitch, Abramowitz, & McKay (2021) | No |
| **Resilience** | | | | |
| Individual Resilience | Connor-Davidson Resilience Scale (CD-RISC-10) | 10 | Campbell-Sills & Stein (2007), adapted from Connor & Davidson (2003) | No |
| Individual Resilience | Mount Sinai Resilience Scale (MSRS) | 24 | DePierro et al. (2023) | No |
| Functional Resilience | Oxford Participation and Activities Questionnaire, Routine Activities (OxPAC-RA) | 14 | Morley et al. (2014); short form: Kelly et al. (2015) | No |
| Posttraumatic Growth | Posttraumatic Growth Inventory (PTGI) | 10 | Tedeschi & Calhoun (1996) | No |

| Construct | Measure Name | # Items | Citation | Modified? |
|---|---|---|---|---|
| Community Resilience | Conjoint Community Resiliency Assessment Measure (CCRAM) | 10 | Leykin, Lahad, Cohen, Goldberg, & Aharonson-Daniel (2013) | No |
| National Resilience | National Resilience Scale | 16 | Kimhi, Eshel, Zysberg, & Leykin (2019); short form (NR-13): Kimhi & Eshel (2016) | Yes – institutional-trust items added |
| **Individual Factors** | | | | |
| Positive Appraisal | Positive Appraisal Style Scale – Content (PASS-Content) | 14 | Petri-Romão et al. (2024) | No |
| Grit | Short Grit Scale (Grit-S) | 8 | Duckworth & Quinn (2009) | No |
| Self-Compassion | Sussex-Oxford Compassion for the Self Scale (SOCS-S) – Modified | 10 | Gu, Baer, Cavanagh, Kuyken, & Strauss (2020) | Yes – top 2 factors per construct retained (10 of 20 original items) |
| Big Five Personality | Ten-Item Personality Inventory (TIPI) | 10 | Gosling, Rentfrow, & Swann (2003) | No |
| Religion | The Duke University Religion Index (DUREL) | 5 | Koenig & Büssing (2010) | Yes – modified for this survey to be culturally appropriate |
| Emotional Regulation | Emotion Regulation Questionnaire – Short Form (ERQ-S) | 6 | Preece, Petrova, Mehta, & Gross (2023) | No |
| Optimism | Life Orientation Test (LOT) | 1 | Scheier & Carver (1985) | Yes – single item only (item #1) |
| **Social Factors** | | | | |
| Loneliness | UCLA Loneliness Scale (3-item) | 3 | Hughes, Waite, Hawkley, & Cacioppo (2004) | No |
| Social Support | Modified Medical Outcomes Study Social Support Survey (mMOS-SS) (mMOS-SS) | 8 | Sherbourne & Stewart (1991) | No |
| Sense of Danger | Sense of Danger Scale | 8 | Adapted from Kimhi & Eshel national resilience research | Yes – translated/adapted for this study |
| **General Mental Health** | | | | |
| Alcohol and Drug Use | ASSIST (Lite) | 11 | WHO ASSIST Working Group (2002) | Yes – limited to past 3 months; tobacco, alcohol, cannabis assessed individually, other drugs grouped |
| Diagnoses / Treatment Status / AI Support | Diagnoses, mental health treatments, use of AI for emotional support | 16 | Investigator-developed | N/A – custom |
| **General Physical Health** | | | | |

| Construct | Measure Name | # Items | Citation | Modified? |
| --- | --- | --- | --- | --- |
| Exercise | International Physical Activity Questionnaire – Short Form (IPAQ-S) | 7 | Craig et al. (2003) | No |
| Eating Habits | Food Noise Questionnaire (FNQ) | 5 | Diktas, Cardel, Foster, et al. (2025) | No |
| Demographics (gender, height, weight, age, preexisting conditions, income, family status, education, residence) | Investigator-constructed demographic items | 12 | Investigator-developed | N/A – standard demographic items |
